# Alignment-Free RoPE-Based Dual-Stream Transformer for PPG-Guided Neonatal ECG Segment Inpainting in the NICU

**DOI:** 10.64898/2026.07.06.26357087

**Authors:** SungLim Choi, Gyeonmin Gu, YunA Kim, Seung Heon Lee, Sang-Il Sim, Yeong Min Jang, Hyunho Kim

**Author notes:** Corresponding author. Email address (Hyunho Kim, MD, PhD). These authors contributed equally to this work (SungLim Choi, Gyeonmin Gu, YunA Kim, Seung Heon Lee, Sang-Il Sim, Yeong Min Jang).

## Abstract

Adhesive electrocardiography (ECG) electrodes used in neonatal intensive care units (NICUs) may cause skin injury in premature infants. Although photoplethysmography (PPG)-based ECG reconstruction has been explored, existing studies have mainly focused on adult data and often rely on direct PPG-to-ECG mapping or artificial signal alignment, which may be unsuitable for neonates with highly variable pulse arrival time (PAT). In this study, we propose an alignment-free RoPE-based dual-stream Transformer for reconstructing missing neonatal ECG segments using concurrent PPG signals and bidirectional ECG context. A total of 52,566 10-second ECG–PPG windows were extracted from 159 NICU patients and split at the patient level to prevent data leakage. The model was designed to learn ECG–PPG temporal coupling without forced synchronization by integrating PPG-derived hemodynamic timing information with lead-specific ECG context. In the primary reconstruction analysis under a 40% random missing condition, the model achieved a Pearson correlation coefficient of 0.96, mean absolute error of 0.04, and root mean square error of 0.07. In stress-test analyses, the model maintained PCC values of at least 0.90 under both 4.0-second continuous block loss and 60% random patch loss. These findings suggest that the proposed framework may serve as a signal imputation module for maintaining ECG monitoring continuity in NICU environments. Prospective validation is required before clinical diagnostic use.

## 1. Introduction

Electrocardiogram (ECG) monitoring is an essential component of vital sign monitoring in the neonatal intensive care unit (NICU). However, conventional ECG monitoring requires adhesive electrodes, which may cause skin injury and increase the risk of infection in neonates with immature skin barriers. Electrode-related skin injury is a particular concern in extremely premature infants because of their immature skin barrier and vulnerability to adhesive-related trauma [1].

Previous studies on ECG reconstruction have mainly focused on adult or general public biosignal datasets. Pinto et al. [2] proposed a modified U-Net-based PPG-to-ECG reconstruction model; however, this approach focuses on full waveform translation from PPG to ECG and does not directly address continuous ECG segment inpainting under irregular long-duration signal loss in neonatal intensive care settings. Other studies have attempted to map photoplethysmography (PPG) to ECG by segmenting individual heartbeats using discrete cosine transform (DCT) or autoencoder-based approaches. However, as highlighted by Abdelgaber et al. [3], many of these models include different beats from the same subject’s record in both training and testing sets, leading to severe subject-dependency and model overfitting. Furthermore, traditional feature extraction relies heavily on the rigid correlation between the R-peak of ECG and the rising curve of the PPG. However, recent clinical evidence suggests that incorporating broader morphological profiles, such as the T-wave or falling curves, holds richer information about hemodynamic transitions and arterial compliance, which are often overlooked in one-to-one mapping frameworks [23]. While deep learning models combining temporal convolutions, LSTMs, or standard attention mechanisms have been developed for PPG-to-ECG conversion [4, 6], their applicability remains limited in extreme missing-signal conditions. Consequently, existing frameworks fail to address the unique physiological characteristics and acute signal misalignment issues of neonates, whose pulse arrival time (PAT) can vary substantially because of immature autonomic regulation.

A more fundamental physiological limitation of prior approaches is that ECG morphology can vary markedly depending on the projection angle of the cardiac axis. Zwanenburg et al. [7] reported that ECG waveform-based markers, including T/QRS-related values, can be influenced by heart-axis orientation. Because of this morphological variability caused by anatomical structure and lead orientation, one-to-one mapping methods that rely solely on PPG signals at a single time point are inherently limited. Although PPG provides useful information for estimating the timing of cardiac cycles, it does not contain the lead-specific morphological profile of the ECG currently being monitored. Therefore, accurate reconstruction of missing ECG waveforms requires not only conversion from PPG but also simultaneous reference to the patient-specific ECG context remaining before and after the missing segment. Such a mechanism is necessary to reconstruct waveforms that are optimized for the corresponding lead orientation.

Preserving ECG morphology is essential for the early diagnosis of congenital heart disease and other cardiac abnormalities in the NICU. However, neonates have distinctive physiological characteristics, including a rapid heart rate of 120–160 beats per minute, narrow QRS duration of less than 60 ms, and frequent body movements, making the direct application of adult algorithms difficult. Previous neonatal monitoring studies have mainly focused on contactless heart-rate estimation using photoplethysmography [8] or ECG signal denoising [9], rather than the morphological reconstruction of missing ECG segments, which is essential when electrode detachment or transient signal loss occurs.

Therefore, the goal of this study is to reconstruct missing ECG segments using concurrent PPG signals and the surrounding ECG context, thereby supporting continuous cardiac monitoring in clinical settings and providing high-quality signal imputation for future data-driven biosignal analysis. In other words, there is a clear need for a method that can morphologically reconstruct transiently missing ECG waveforms using non-invasive physiological signals and bidirectional ECG context, while reducing the risk of skin injury associated with adhesive ECG electrodes.

In this study, we propose an ECG segment-level reconstruction framework using concurrent PPG signals and bidirectional ECG context. By introducing neonatal-specific preprocessing and an alignment-free dual-stream Transformer, the proposed method is designed to learn physiological PAT-related temporal relationships without forced signal alignment and to preserve waveform continuity and morphology even under severe missing-signal conditions.

## 2. Methods

### 2.1. Study Population and Data Splitting

This study was conducted with the approval of the Institutional Review Board (IRB) of Jeonbuk National University Hospital (IRB No. 2025-12-052-001). Physiological waveform data recorded between January 2024 and December 2025 were obtained from 412 infants admitted to the neonatal intensive care unit (NICU) of Jeonbuk National University Hospital. To record and extract high-resolution, time-synchronized physiological data from patient monitors without omissions, we used NICU waveform data recorded by Vital Recorder [10], an automatic physiological data recording program. Applying the inclusion criteria targeting premature infants with a gestational age of less than 37 weeks and a birth weight of less than 2,500 g, a total of 203 eligible subjects were selected.

Subsequently, to ensure high-quality signals for model training, specific exclusion criteria were additionally applied. The exclusion criteria were as follows:

First, cases without paired, time-synchronized PPG and ECG signals were excluded.

Second, mechanical flatline segments with no amplitude variations caused by sensor detachment, as well as segments where the signal was continuously lost for 5 seconds (50%) or more within a 10-second analysis window, were excluded.

Third, invalid waveform segments in which physiological reference points, including the ECG QRS complex and PPG systolic peak, could not be reliably identified because of severe motion artifacts or non-physiological device-level desynchronization were excluded from the analysis.

Due to such excessive noise and signal omissions, data from 44 additional infants were excluded. The final analysis cohort comprised data from 159 infants (Figure 1).

**Figure 1:**
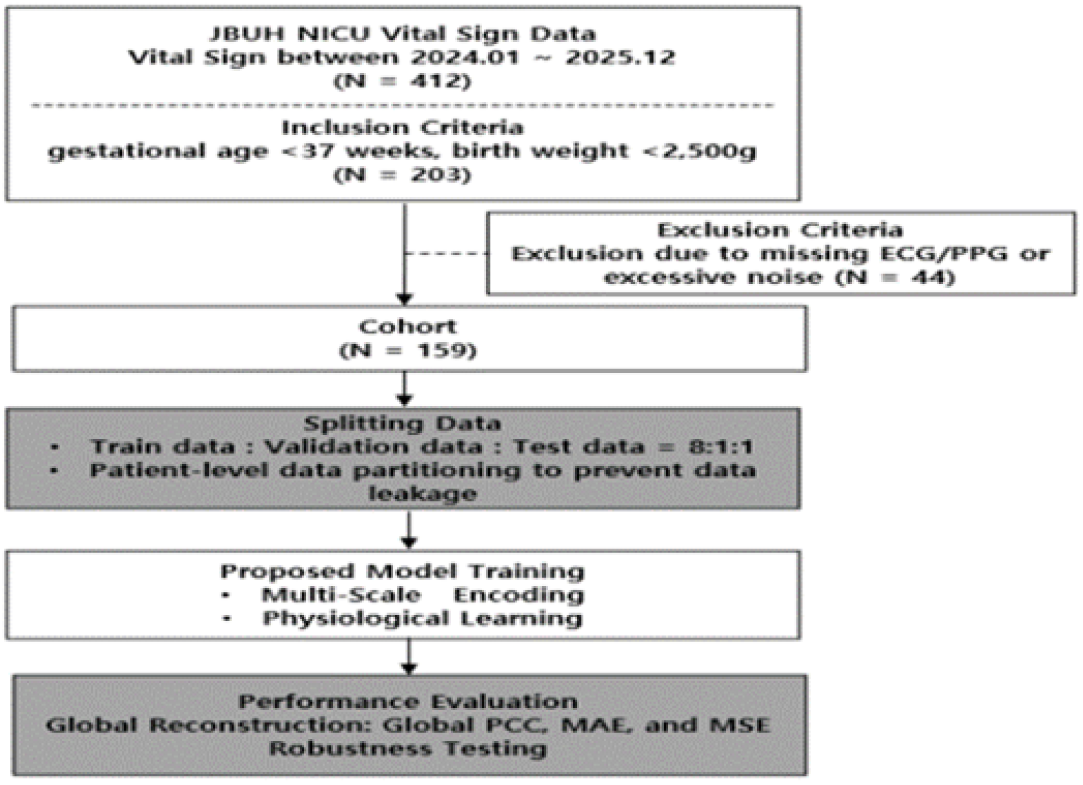
Study flowchart.

To ensure objectivity in the model evaluation and mitigate data leakage, the constructed dataset was strictly partitioned at the patient level into training, validation, and test sets at a ratio of 8:1:1, respectively. This patient-level split was used to reduce the risk of subject-dependent performance inflation and dataset-shift-related overestimation [18].

### 2.2. Data Preprocessing

The collected and synchronized photoplethysmography (PPG) and electrocardiogram (ECG) signals were segmented into 10-second windows for time-series analysis and used as model inputs. To ensure high signal quality, invalid segments, including mechanical flatline patterns caused by sensor detachment and segments with excessive signal loss, were strictly excluded during the preprocessing stage.

Unlike adult ECGs, neonatal ECGs are characterized by steep R-peaks and narrow QRS complexes. Therefore, the uniform application of conventional general-purpose filters may lead to morphological distortion of key waveform components (Figure 2). To address this issue, we applied a premature-infant-specific adaptive hybrid preprocessing pipeline. This pipeline adopted a dual low-pass filtering strategy, in which different frequency bands were selectively applied to the relatively smooth P- and T- wave regions and to the QRS complex regions, where high-frequency components are concentrated. By applying component-specific low-pass filtering, the proposed preprocessing pipeline was designed to preserve neonatal ECG morphology while improving signal quality (Figure 3).

**Figure 2:**
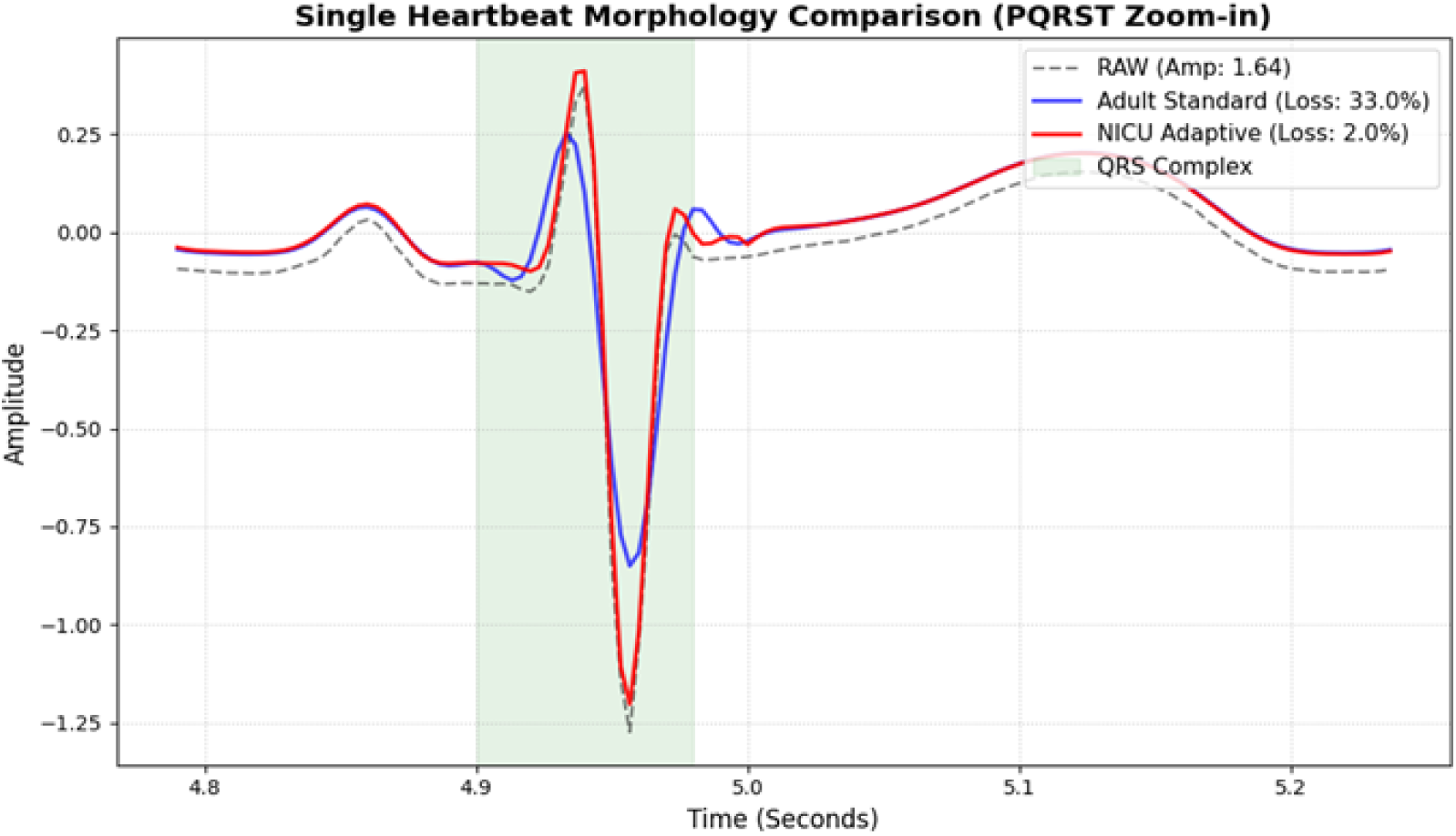
NICU adaptive filter.

**Figure 3:**
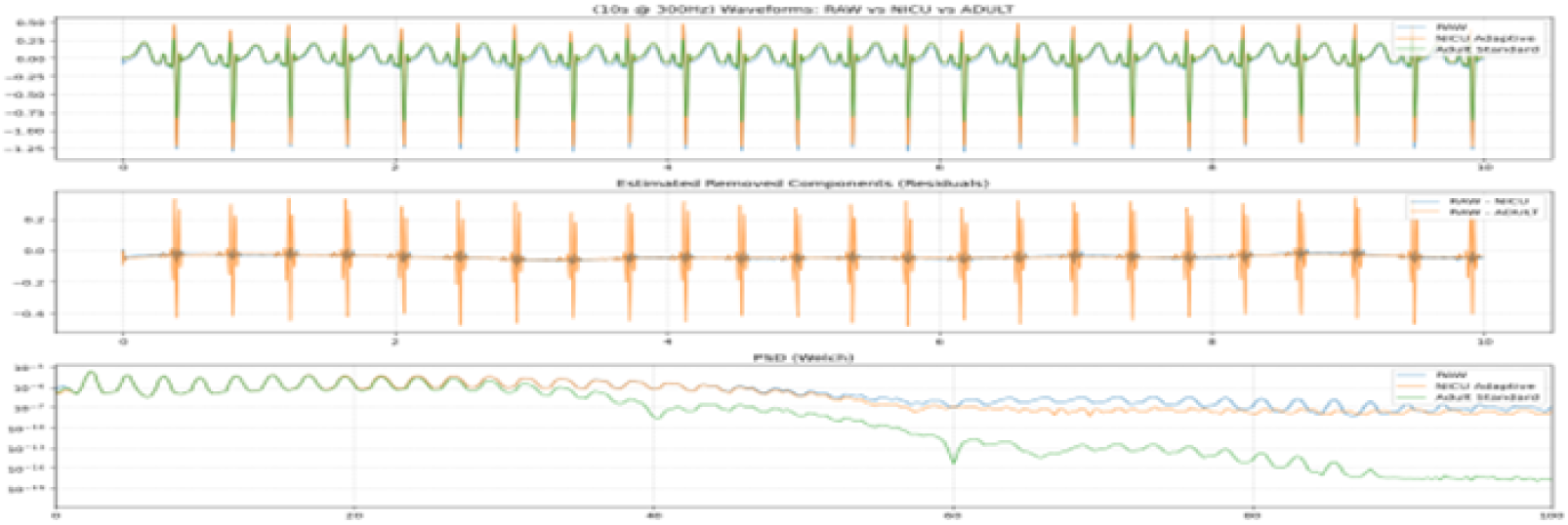
Hybrid preprocessing pipeline.

To remove baseline wander and powerline interference from the electrocardiogram (ECG) signals, a 0.7 Hz high-pass filter and a 60 Hz notch filter were first applied, following standard ECG preprocessing principles for suppressing low-frequency drift and mains interference [30]. The dual low-pass filtering (Dual LPF) strategy was introduced because different ECG components have distinct frequency characteristics; the T wave is mainly distributed in the low-frequency range, whereas the P wave and QRS complex contain relatively higher-frequency components [31]. Accordingly, for the relatively smooth P- and T-wave regions, a 25–40 Hz low-pass filter was applied to suppress residual noise while preserving waveform smoothness. In contrast, for the QRS complex regions, where higher-frequency components are concentrated, a 40–70 Hz low-pass filter was applied to reduce morphological distortion of the narrow and sharp peaks. This component-specific filtering strategy was designed to preserve the sharp and narrow morphology of neonatal ECG waveforms [32].

After generating signals optimized for each frequency band, the two filtered signals were cross-merged according to ECG component regions. Finally, Savitzky–Golay smoothing was applied to the hybrid ECG signal using an 11-sample window length, corresponding to approximately 36.7 ms at 300 Hz, and a third-order polynomial to preserve local signal trends while further improving signal quality [11]. This preprocessing strategy was designed to enhance signal quality while minimizing distortion of clinically important ECG waveform components. The component-specific ECG filtering was performed during offline signal preprocessing before artificial masking was applied. During model training and evaluation, the artificially masked ECG samples were not provided to the model, and the binary mask explicitly indicated the unavailable regions. Therefore, the model input did not contain waveform information from the target missing segments.

For the photoplethysmography (PPG) signals used as model input, a 0.5–18.0 Hz band-pass filter was applied, reflecting the physiological characteristic that PPG information is concentrated in a relatively lower frequency range than ECG. This process reduced respiratory baseline wander below 0.5 Hz and high-frequency noise above 18.0 Hz, including motion artifacts and optical sensor noise, thereby obtaining pulse waveforms suitable for model training (Figure 4).

**Figure 4:**
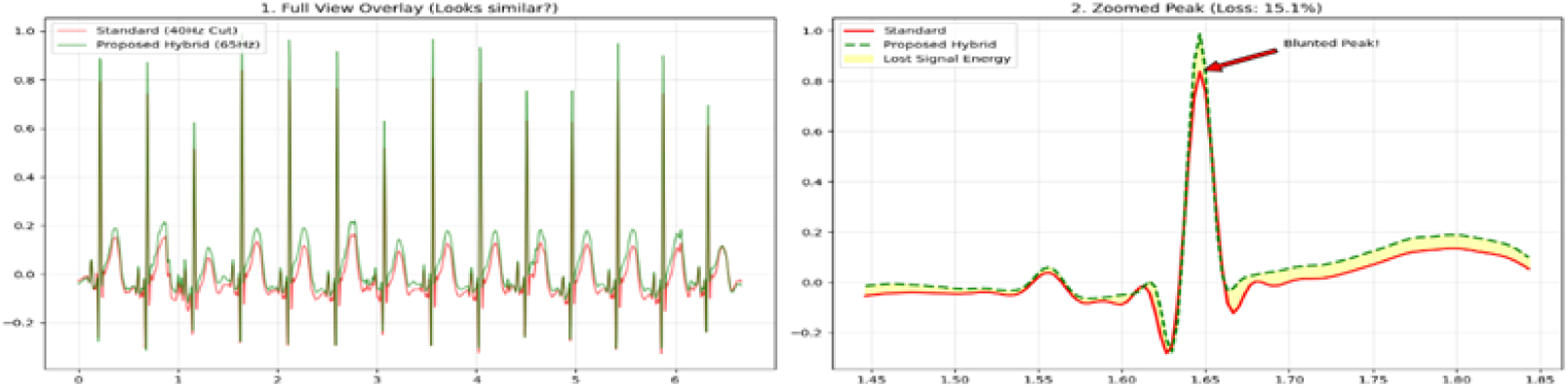
Zoomed ECG waveform after preprocessing.

A 10-second sample of the training data set is presented, ultimately constructed through the previously proposed hybrid preprocessing pipeline (Figure 5). The red graph at the top represents the electrocardiogram (ECG) signal, which serves as the reconstruction target for the model, while the blue graph at the bottom displays the photoplethysmography (PPG) signal used as the model’s input. In particular, within the ECG signal, the sharp peaks of the QRS complex, along with the fine morphological characteristics of the P and T waves, which are essential for clinical diagnosis, are preserved without any distortion. Furthermore, the physiological Pulse Arrival Time (PAT) delay in the time required for blood to reach the peripheral vessels after ventricular contraction is naturally maintained between the two waveforms. This provides an optimal foundation for the proposed alignment-free architecture to autonomously learn temporal delay characteristics without requiring any artificial alignment manipulation.

**Figure 5:**
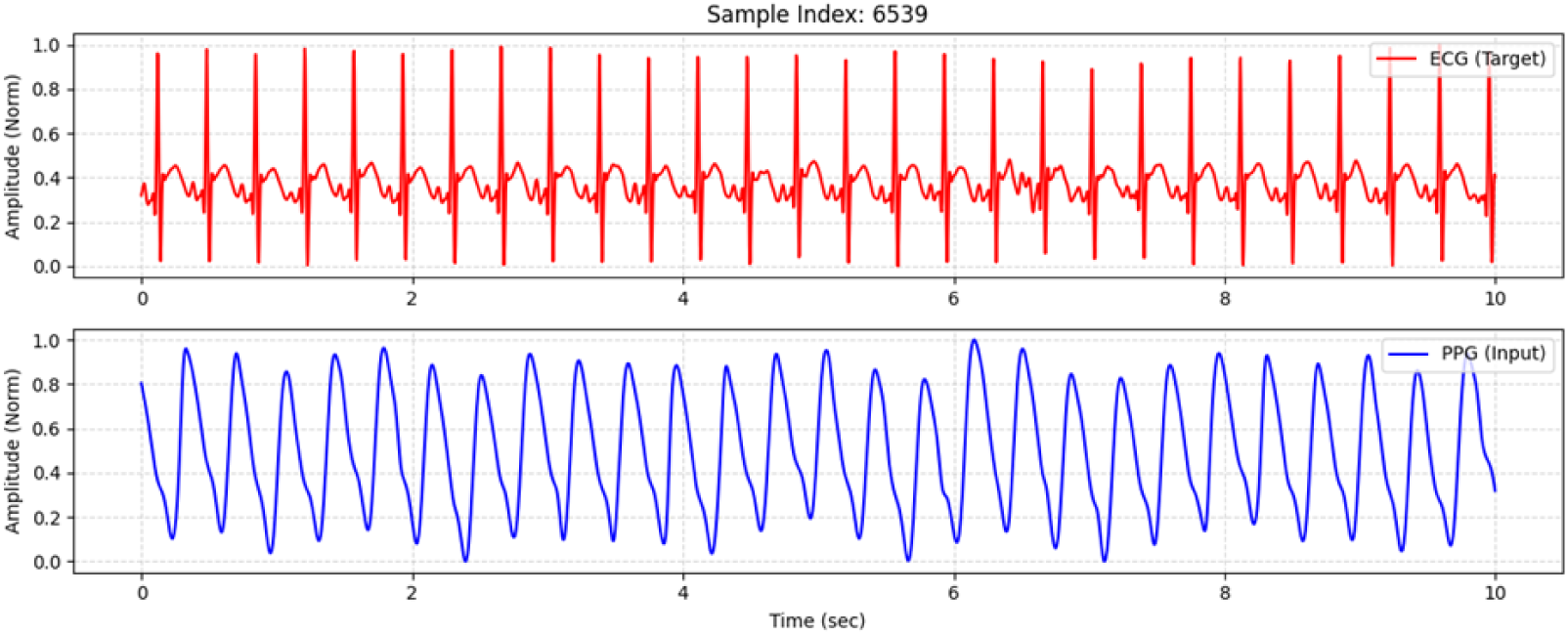
Output of the adaptive hybrid preprocessing pipeline.

### 2.3. Proposed Model Architecture

In this study, we propose a RoPE-based alignment-free dual-stream Transformer architecture to perform morphological partial reconstruction of long missing-signal segments lasting several seconds. The proposed architecture is designed to capture long-range dependencies in time-series data and model relative positional relationships. Conventional convolutional neural network (CNN)- or long short-term memory (LSTM)-based models may suffer from information bottlenecks or gradient vanishing when processing long sequences of 3,000 time steps, corresponding to 10 seconds at 300 Hz. In contrast, the proposed model uses a self-attention mechanism to directly connect the global context before and after the missing segment, thereby preserving waveform continuity.

A key feature of the proposed model is its multi-view input representation and multi-scale tokenization strategy. The encoder was designed to incorporate both the raw PPG signal and its first derivative, dPPG/dt, to reflect the dynamic rate of change in blood flow. Because different ECG components have distinct durations and frequency characteristics, the reference PPG signal was segmented using three different window sizes of 50, 100, and 200 samples. The short window of 50 samples captures subtle inflection points in the pulse waveform and helps guide the timing of QRS peak occurrence, whereas the long window of 200 samples captures the overall cardiac cycle and contributes to maintaining the morphology of smoother T-wave components and stable heart rhythm, including RR intervals.

The rationale for adopting rotary position embedding (RoPE), instead of the absolute positional encoding used in the original Transformer [12], was to accommodate both the periodicity and variability of biosignals. RoPE encodes relative positional dependencies through vector rotation and can capture token-to-token positional relationships in long sequences [13]. Although ECG is a periodic waveform, the absolute positions of its components can vary because of heart rate variability (HRV), which limits the effectiveness of fixed positional encoding. In contrast, RoPE represents relative distances between tokens as rotation angles, allowing the model to capture relationships among P-QRS-T components regardless of their absolute positions. This property is particularly useful for preserving contextual relationships between distant tokens under long missing-signal conditions, such as gaps lasting 4.0 seconds or longer.

Based on this property, the proposed model applies relative rotational encoding to learn positional relationships between tokens. Specifically, RoPE was applied to the self-attention layers of both the encoder and decoder to enhance internal contextual modeling. In contrast, RoPE was intentionally excluded from the cross-attention module. This design was intended to allow the model to learn the intrinsic physiological time delay between PPG and ECG, namely pulse arrival time (PAT), without relying on forced peak alignment. In an ablation analysis, applying RoPE to the cross-attention module decreased the Pearson correlation coefficient (PCC) from 0.96 to 0.93. This result suggests that the alignment-free design of the proposed model better accommodates the substantial PAT variability observed in neonates and contributes to improved morphological reconstruction performance.

In the final reconstruction stage, ECG tokens generated by the decoder were reassembled into a continuous signal using a fold operation. Overlapping signal patches were combined through weighted averaging, which reduced boundary artifacts such as abrupt discontinuities or spikes between adjacent patches. This process enabled the model to generate smooth and physiologically plausible ECG waveforms (Figure 6).

**Figure 6:**
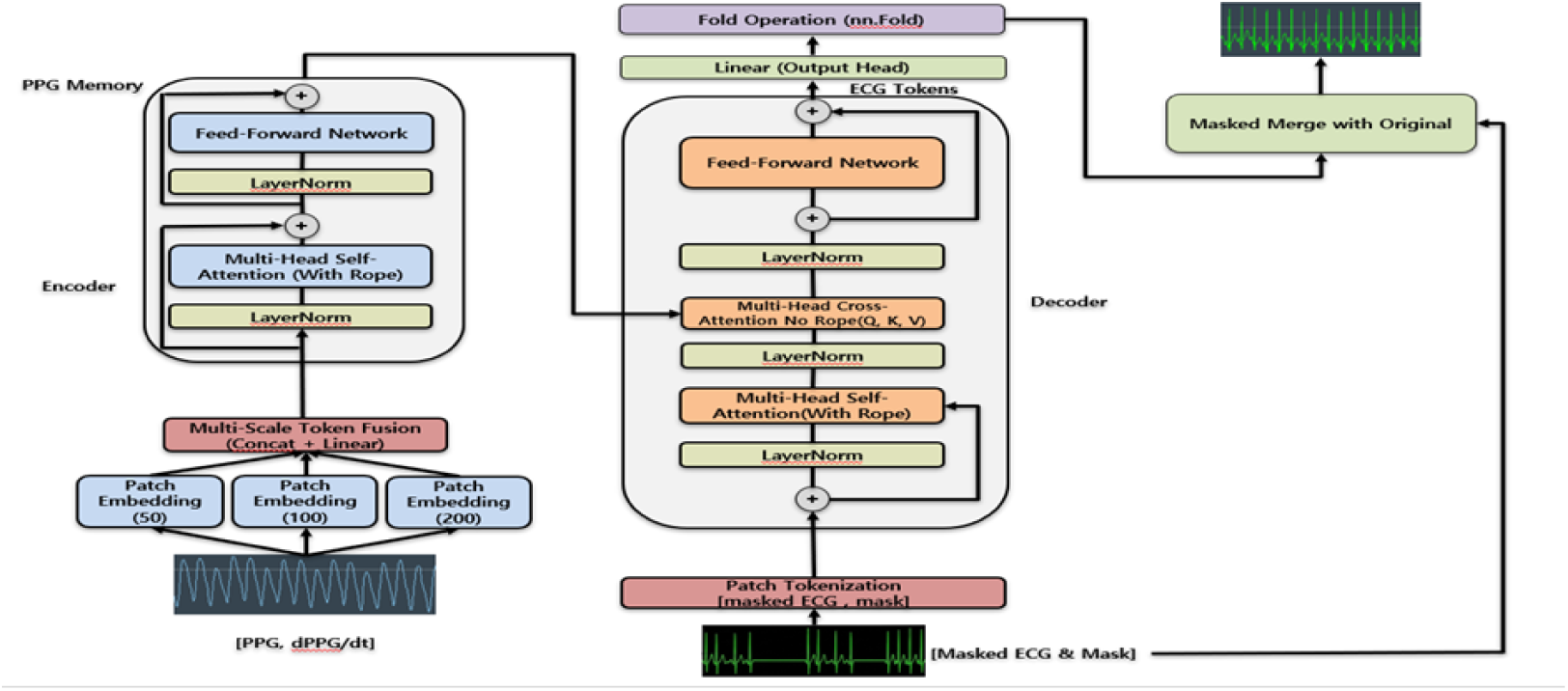
Model architecture.

### 2.4. Implementation Details

All experiments were implemented using PyTorch. The ECG and PPG signals were sampled at 300 Hz, and each input window had a duration of 10 seconds, corresponding to 3,000 samples. The PPG signal was normalized to the range of [*−*1, 1] using window-wise min–max scaling. For the masked ECG input, scaling parameters were computed only from the observed, non-masked ECG samples within each window and then applied to the corresponding masked ECG input. This procedure was used to avoid using amplitude information from the target missing segment during model input preparation. The input consisted of three channels: the PPG signal, the masked ECG signal, and the binary mask indicating missing ECG samples.

The proposed model used a model dimension of 256 with eight attention heads. The PPG encoder consisted of three RoPE-based Transformer encoder layers, whereas the ECG decoder consisted of eight RoPE-based Transformer decoder layers. The dropout rate was set to 0.1. For the PPG encoder, multi-scale tokenization was performed using patch sizes of 50, 100, and 200 samples with a stride of 20 samples. The ECG decoder used patches of 100 samples with a stride of 20 samples. RoPE was applied to the self-attention layers of both the encoder and decoder, whereas conventional cross-attention without RoPE was used between the ECG decoder and PPG encoder to preserve the alignment-free ECG–PPG temporal learning scheme.

The model was trained for up to 100 epochs using the AdamW optimizer with an initial learning rate of 8 *×* 10^−5^ and a batch size of 16. Early stopping was applied with a patience of 15 epochs based on validation loss. Mixed-precision training was performed using automatic mixed precision and gradient scaling. During training, random patch masking with a masking ratio of 40% was used, and continuous block masking was applied with a probability of 40%. The block-mask duration was gradually increased during training from 0.5–1.5 seconds in the early stage to 4.0–5.0 seconds in the final stage. For reproducible evaluation, fixed random seeds were used for validation and test masking.

### 2.5. Performance Evaluation

To objectively evaluate the reconstruction performance of the proposed model, we constructed two artificial masking conditions that reflect different types of signal loss commonly encountered in clinical environments. First, to simulate intermittent signal loss caused by motion artifacts, a random patch masking condition was generated by masking short patches at multiple randomly selected time points within each 10-second ECG Lead I window, resulting in 40% loss of the total signal length. Second, to evaluate long- duration signal interruption caused by electrode detachment, a continuous block masking condition was designed by randomly selecting a starting point within the window and masking a continuous segment ranging from 1.0 to 4.0 seconds.

The model was trained to infer the missing ECG segments by referring to the concurrent complete PPG signal. The originally measured ECG waveform before masking was used as the ground truth for calculating reconstruction accuracy. To reproduce the physiological characteristics of the ECG waveform during training, an integrated reconstruction loss function was applied. This loss combined amplitude error, phase similarity, and temporal continuity by incorporating L1/MSE loss, Pearson correlation coefficient (PCC)-based loss, and difference loss between the predicted and ground-truth waveforms (Figure 7).

**Figure 7:**
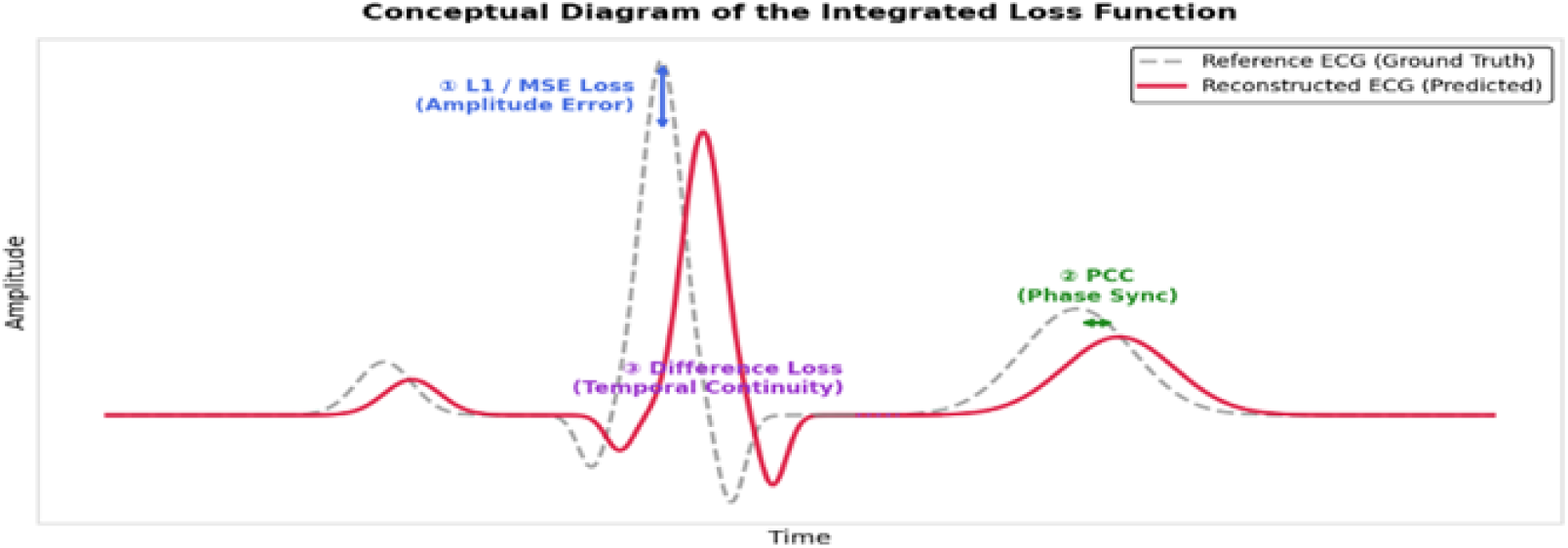
Integrated loss function.

Furthermore, during the evaluation phase, reconstruction performance was assessed from multiple perspectives, reflecting recent clinical evidence that rhythm evaluation may be difficult when relying solely on amplitude- based errors such as mean absolute error (MAE) [14]. Therefore, global MAE and global Pearson correlation coefficient (PCC) were calculated within the masked regions to quantify the absolute error and morphological similarity between the reconstructed and original ECG waveforms. In addition, component-wise PCC was introduced to evaluate the morphological preservation of clinically important ECG components, including the P wave, QRS complex, and T wave, thereby enabling a more detailed assessment of agreement with the ground-truth waveform (Figure 8).

**Figure 8:**
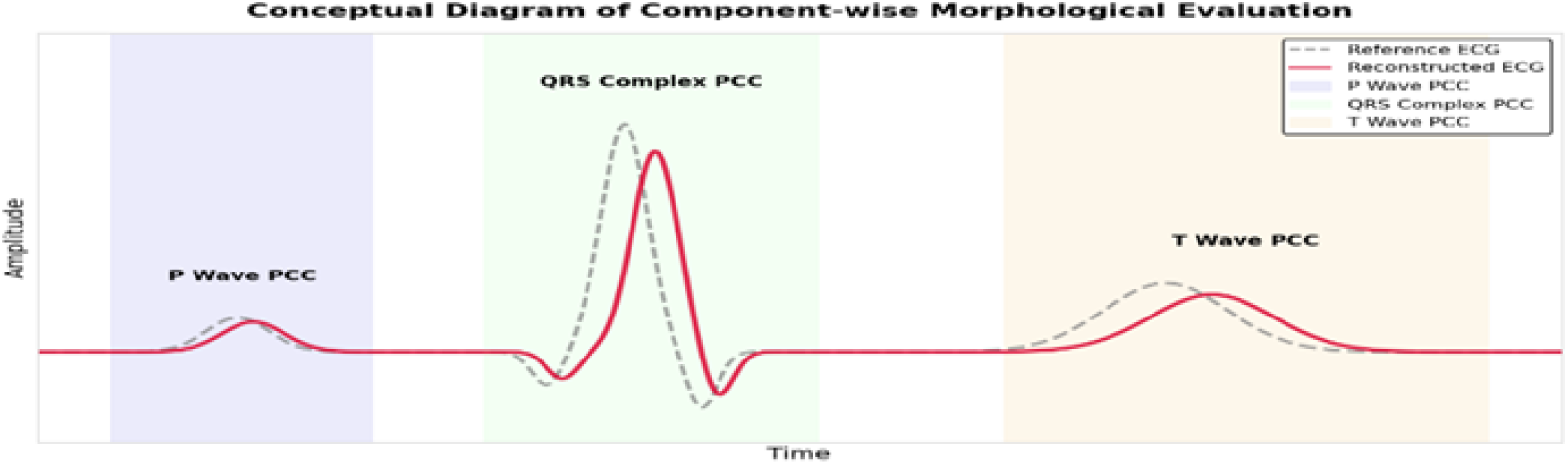
Component-wise PCC.

## 3. Results

### 3.1. Cohort Characteristics and Training Data Construction

A total of 52,566 10-second ECG–PPG windows were constructed from the final cohort of 159 NICU patients. To reduce patient-level imbalance during model training, the maximum number of extracted windows per patient was capped at 600. The dataset was then split at the patient level into training, validation, and test sets at a ratio of 8:1:1. The final dataset consisted of 42,126 training windows from 127 patients, 4,687 validation windows from 16 patients, and 5,753 test windows from 16 patients.

Clinical characteristics were summarized at the patient level and are presented as mean *±* standard deviation or number with percentage, as appropriate. The overall cohort had a gestational age of 31.9 *±* 3.0 weeks and a birth weight of 1749.4 *±* 499.3 g. To assess potential imbalance after patient-level splitting, the Shapiro–Wilk test was first performed for gestational age and birth weight. Because some variables did not satisfy the normality assumption, the non-parametric Kruskal–Wallis test was subsequently used. No statistically significant differences were observed among the training, validation, and test groups in gestational age (*p* = 0.1235) or birth weight (*p* = 0.1595), suggesting that the patient-level split resulted in comparable clinical characteristics across the three groups.

In addition, ECG–PPG temporal delay was calculated at the window level. The unaligned ECG–PPG windows showed an average delay of 172.9 *±* 72.1 ms and were used as input data without artificial phase alignment. This setting allowed the model to learn temporal delays that may occur in real NICU monitoring environments (Table 1).

**Table 1:** Patient-level clinical characteristics according to dataset division.

| Characteristic | All<br>(N=159) | Train<br>(N=127) | Val<br>(N=16) | Test<br>(N=16) | P value |
| --- | --- | --- | --- | --- | --- |
| Male | 95 (59.7%) | 74 (58.3%) | 10 (62.5%) | 11 (68.8%) | - |
| GA (weeks) | $31.9 \pm 3.0$ | $32.1 \pm 2.9$ | $31.8 \pm 2.6$ | $30.5 \pm 3.9$ | 0.12 |
| Birth weight (g) | $1749.4 \pm 499.3$ | $1785.3 \pm 495.0$ | $1653.1 \pm 466.4$ | $1560.3 \pm 539.2$ | 0.16 |
| RDS | 68 (42.8%) | 51 (40.2%) | 11 (68.8%) | 6 (37.5%) | - |

### 3.2. Reconstruction Performance and Robustness Evaluation

#### 3.2.1. Comparison of Model Performance

All baseline models were trained and evaluated using the same patient-level data split, preprocessing pipeline, input window length, masking strategy, and evaluation metrics as the proposed model. The same training, validation, and test sets were used for all model comparisons to ensure a fair comparison.

The performance of the proposed RoPE-based Dual-Stream Transformer was evaluated under a random 40% missing condition. The proposed model achieved the best overall performance, with a mean PCC of 0.96 *±* 0.05, MAE of 0.04 *±* 0.02, and RMSE of 0.07 *±* 0.03. These primary reconstruction metrics were calculated within the masked ECG regions to directly quantify the accuracy of missing-segment reconstruction. To assess window-level reconstruction performance under the test condition, these descriptive statistics were calculated using 5,753 windows from the test dataset.

However, for statistical hypothesis testing, a strict patient-level analysis was performed to reduce the risk of type I error caused by dependency among multiple windows from the same patient. For each of the final 16 test patients who passed the signal quality assessment, the mean performance of each model was calculated. Normality testing showed that the normality assumption was violated (*p <* 0.05). Therefore, the non-parametric Friedman test was performed, followed by post hoc Wilcoxon signed-rank tests.

The proposed model showed superior reconstruction performance compared with the baseline models, including WNet-BiLSTM (PCC 0.86) [4], Performer (PCC 0.91) [5], 1D U-Net (PCC 0.54) [16], and TCN (PCC 0.88) [17]. Patient-level statistical testing showed significant improvements in PCC and MAE compared with the baseline models (*p <* 0.001, Table 2). Notably, the proposed model also achieved a lower reconstruction error than Performer, which showed the highest PCC among the baseline models. These findings suggest that the proposed model more accurately reconstructs the morphological characteristics of neonatal ECG signals under the tested missing-signal condition.

**Table 2:** Model comparison performance under random 40% missing condition.

| Model | PCC | MAE | RMSE |
| --- | --- | --- | --- |
| RoPE-based Dual-Stream Transformer | $0.96 \pm 0.05^*$ | $0.04 \pm 0.02^*$ | $0.07 \pm 0.03$ |
| 1D U-Net | $0.54 \pm 0.12$ | $0.24 \pm 0.10$ | $0.31 \pm 0.10$ |
| TCN | $0.88 \pm 0.11$ | $0.08 \pm 0.03$ | $0.12 \pm 0.04$ |
| WNet-BiLSTM | $0.86 \pm 0.10$ | $0.08 \pm 0.03$ | $0.14 \pm 0.04$ |
| Performer | $0.91 \pm 0.08$ | $0.06 \pm 0.02$ | $0.10 \pm 0.03$ |

\* $p < 0.001$ compared with baseline models using post hoc Wilcoxon signed-rank tests after the Friedman test.

In the ablation study, blocking the PPG input led to a marked decrease in PCC from 0.96 to 0.59. The MAE also increased from 0.04 to 0.11, indicating greater absolute reconstruction error. These consistent performance degradations suggest that PPG provides essential complementary information for reconstructing missing ECG segments, particularly hemodynamic timing and rhythm-related cues (Figure 9).

**Figure 9:**
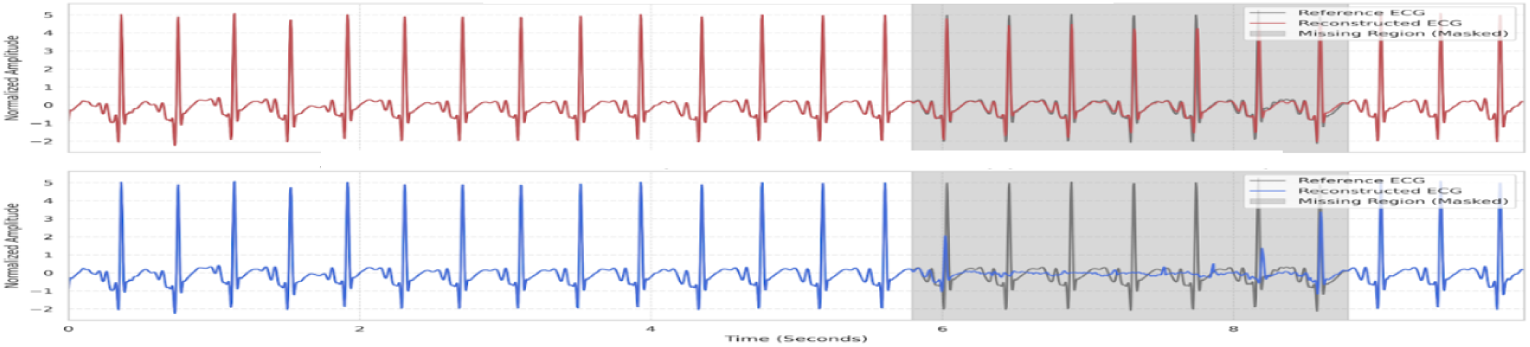
Ablation study.

Specifically, the attention map analysis showed that, during R-peak reconstruction, 72.0% of the attention weights were concentrated around the foot/onset region of the subsequent PPG pulse. This pattern is physiologically plausible because the temporal interval between the ECG R-peak and fiducial points of the peripheral PPG waveform, including the PPG foot/onset, has been commonly used to estimate pulse arrival time (PAT) or PAT-like timing [26, 27]. The learned ECG–PPG temporal offset of 143.2 *±* 88.1 ms may indicate that the model utilized PAT-like temporal information without explicit temporal supervision. In addition, the relatively large variability of this learned delay may be related to beat-to-beat variability in ECG–PPG coupling in premature infants, whose autonomic regulation and heart-rate dynamics are still maturing [28]. Time-based ECG–PPG parameters have also been investigated in preterm infants using pulse oximeter and ECG waveforms, supporting the relevance of this temporal relationship in neonatal monitoring [29]. However, attention weights should be interpreted as exploratory evidence rather than direct causal explanations of model behavior (Figure 10).

**Figure 10:**
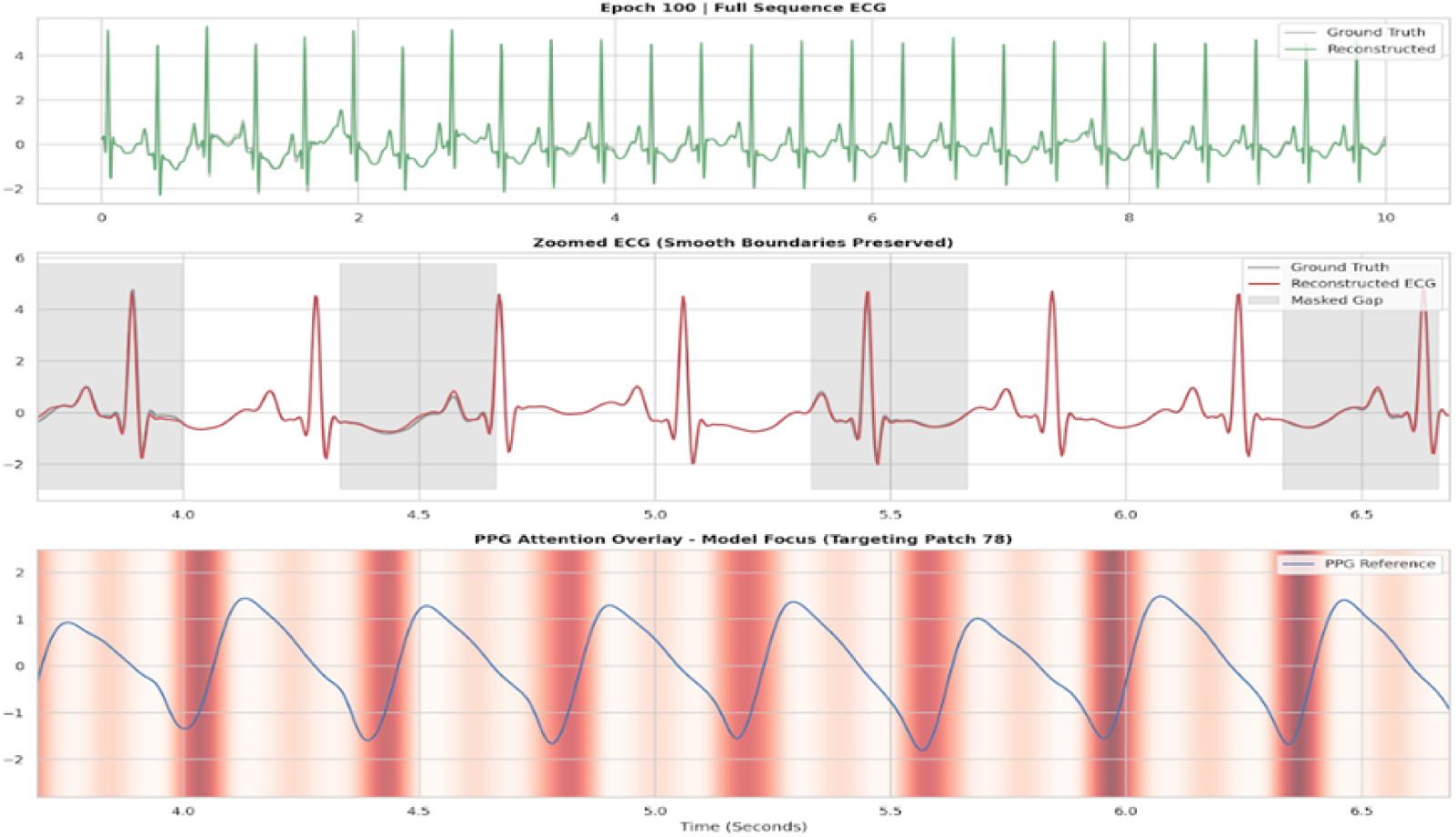
Attention map.

#### 3.2.2. Morphological Preservation and Robustness Analysis

The proposed model preserved the morphology of major ECG components, including the QRS complex (PCC 0.96), T wave (PCC 0.91), and P wave (PCC 0.92), supporting the morphological consistency of the reconstructed neonatal ECG waveforms. In particular, single reconstructed heartbeats, including P-QRS-T complexes, were extracted and visualized using t-distributed stochastic neighbor embedding (t-SNE) to assess their morphological distribution. The t-SNE visualization provided exploratory evidence that reconstructed ECG beats occupied a similar morphological embedding space to the corresponding ground-truth beats (Figure 11).

**Figure 11:**
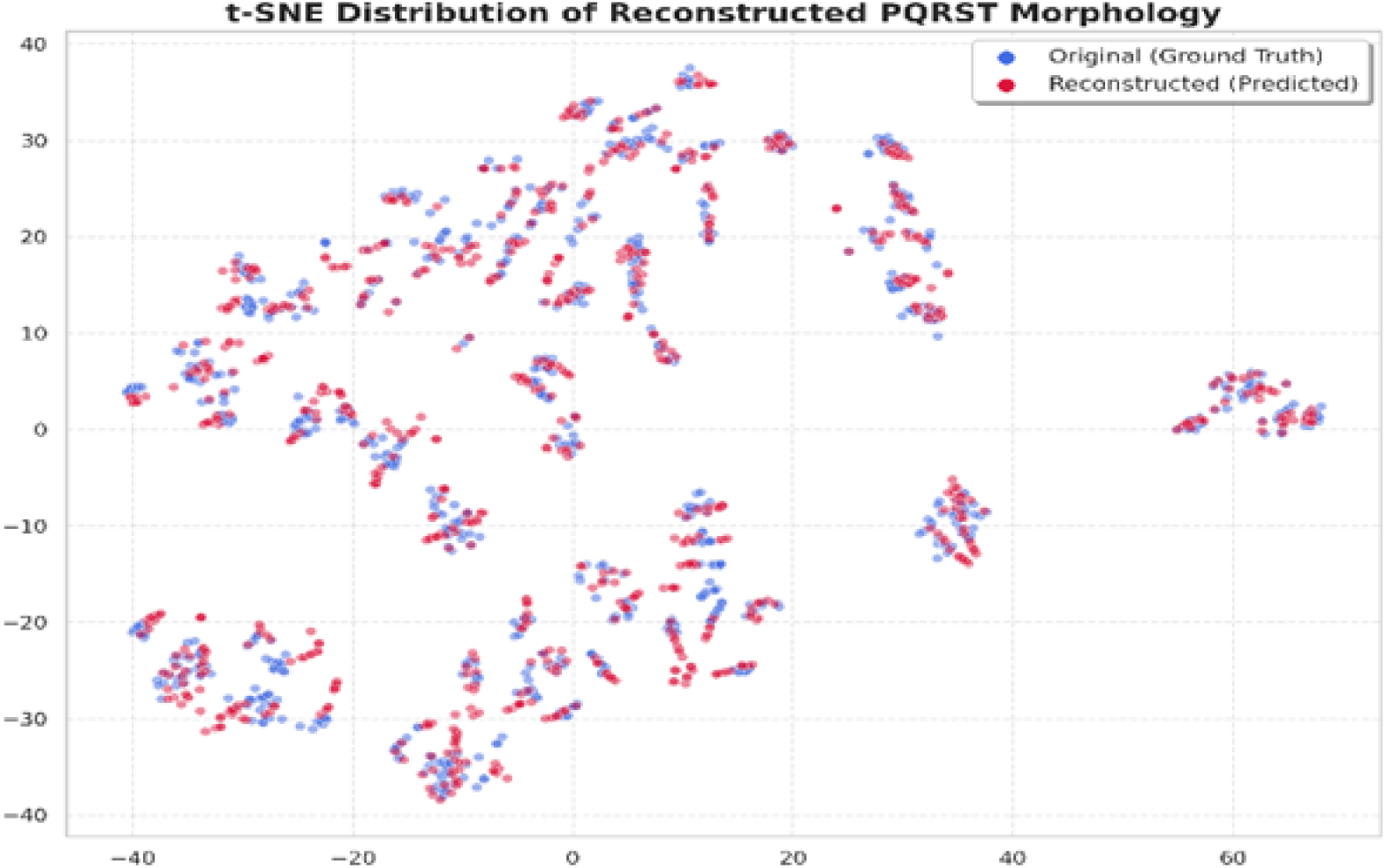
t-SNE distribution of reconstructed PQRST morphology.

In the robustness evaluation simulating sensor detachment, the performance degradation threshold was carefully analyzed. The proposed model maintained a Global PCC of 0.90 up to a 4.0-second continuous missing segment, which was the longest block-loss condition tested in this study. Furthermore, even under this severe signal-loss condition, the model preserved overall rhythm inference capability, suggesting its potential applicability for maintaining ECG monitoring continuity in NICU environments (Table 3, Figure 12).

**Figure 12:**
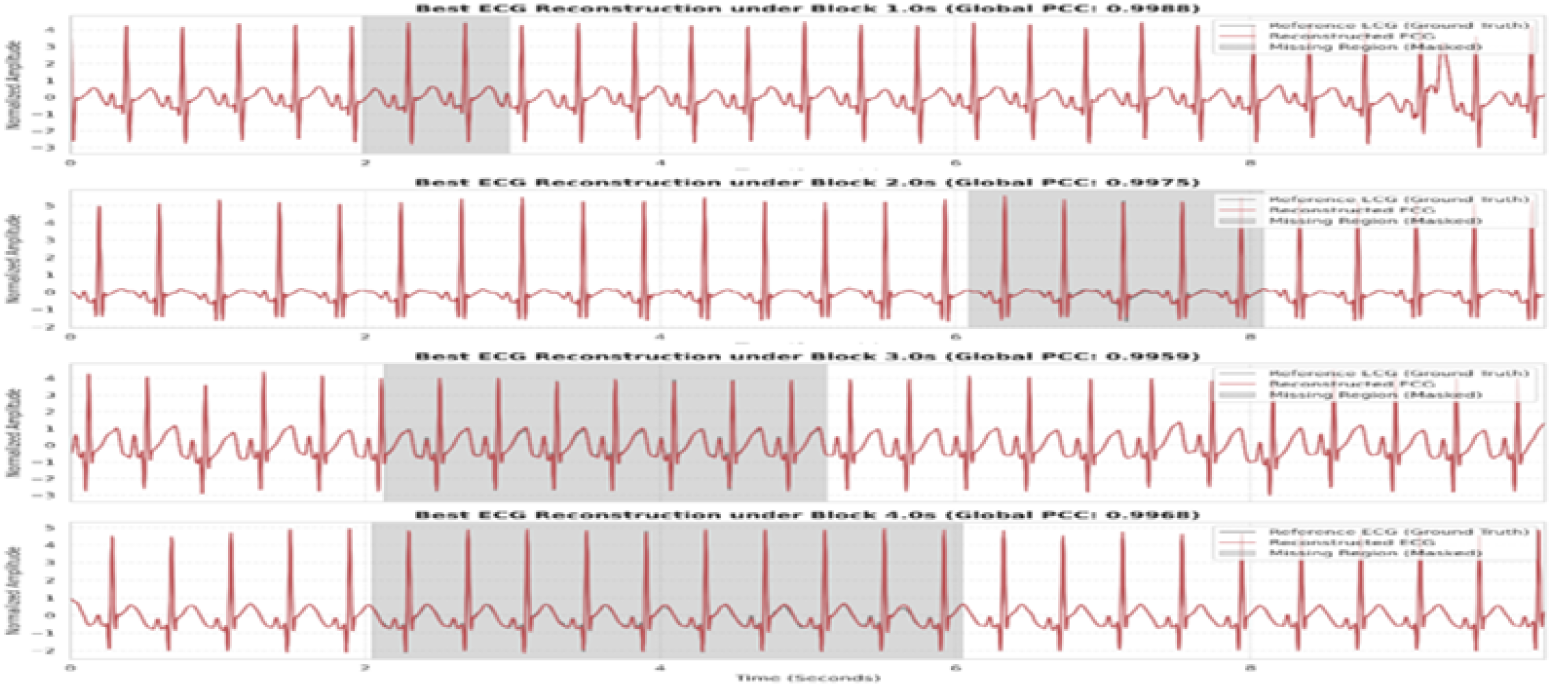
Testing block length.

**Table 3:** Reconstruction performance under continuous block loss.

| Block length (s) | Global PCC | QRS PCC |
| --- | --- | --- |
| 1.0 | $0.96 \pm 0.07$ | $0.96 \pm 0.10$ |
| 2.0 | $0.94 \pm 0.08$ | $0.94 \pm 0.13$ |
| 3.0 | $0.92 \pm 0.10$ | $0.93 \pm 0.15$ |
| 4.0 | $0.90 \pm 0.11$ | $0.90 \pm 0.20$ |

For random patch loss, the performance degradation threshold was carefully analyzed. The proposed model maintained robust reconstruction performance even when 60% of the ECG samples were randomly masked, achieving a Global PCC of 0.93 and a QRS PCC of 0.94. In contrast, performance decreased under the 80% random patch loss condition (Global PCC 0.85), likely because the available ECG context and PPG-derived physiological cues became insufficient for reliable reconstruction.

Because the model was trained with a 40% random patch masking ratio, the 60% and 80% random patch loss conditions were used as stress-test settings that exceeded the nominal random masking ratio used during training. Although the model was also exposed to continuous block masking during curriculum training, random patch loss introduces a different pattern of distributed signal removal. Therefore, the preserved performance under the 60% random patch condition suggests that the model did not simply memorize a specific masking pattern, but learned robust temporal relationships between the PPG signal and the surrounding ECG context.

Based on these findings, we identified 60% random patch loss as the maximum tested random missing condition under which the model maintained reliable reconstruction performance. This result supports the potential applicability of the proposed framework in NICU environments where frequent and unpredictable motion artifacts may cause intermittent ECG signal loss (Table 4, Figure 13).

**Figure 13:**
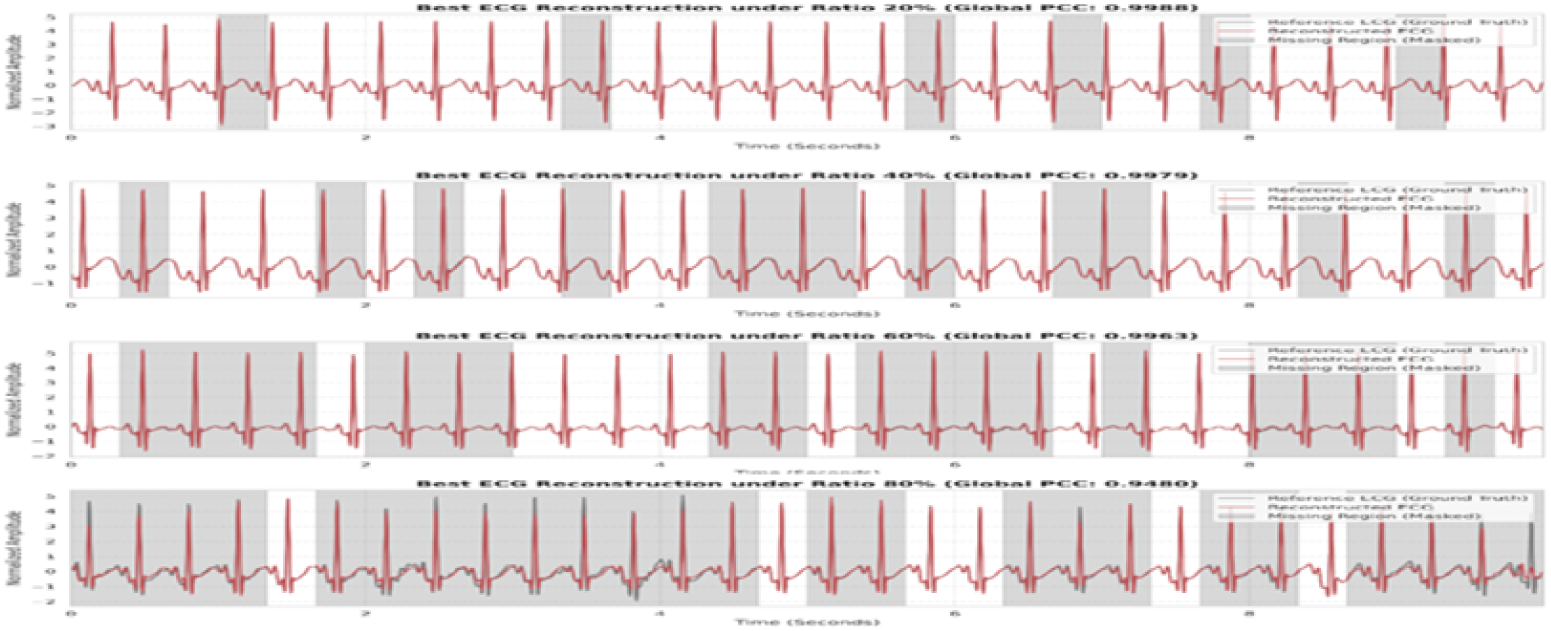
Testing missing ratio.

**Table 4:** Reconstruction performance under random patch loss.

| Missing ratio (%) | Global PCC | QRS PCC |
| --- | --- | --- |
| 20 | $0.96 \pm 0.05$ | $0.97 \pm 0.11$ |
| 40 | $0.95 \pm 0.05$ | $0.96 \pm 0.11$ |
| 60 | $0.93 \pm 0.07$ | $0.94 \pm 0.17$ |
| 80 | $0.85 \pm 0.13$ | $0.83 \pm 0.31$ |

## 4. Discussion

### 4.1. Principal Findings

In this study, we developed an alignment-free RoPE-based dual-stream Transformer to reconstruct missing electrocardiogram (ECG) segments using non-invasive photoplethysmography (PPG) and bidirectional ECG context. The proposed framework was designed to address a clinically important limitation of conventional neonatal intensive care unit (NICU) monitoring, namely the dependence on adhesive ECG electrodes, which may contribute to medical adhesive-related skin injuries (MARSI) in premature infants [1, 15]. Under a random 40% missing condition, the proposed model achieved high morphological reconstruction performance, with a Pearson correlation coefficient (PCC) of 0.96, mean absolute error (MAE) of 0.04, and root mean square error (RMSE) of 0.07. In addition, the model maintained robust reconstruction performance under clinically challenging missing-signal scenarios, including 4.0-second continuous block loss and 60% random patch loss. These findings suggest that the proposed model did not merely learn a specific masking pattern, but captured physiologically meaningful temporal relationships between PPG and ECG.

### 4.2. Core Conceptual Shift: ECG Segment Inpainting

The most critical conceptual contribution of this work is that this study should be interpreted as ECG segment-level inpainting rather than full PPG-to-ECG synthesis. Conventional PPG-to-ECG studies attempt to generate an entire ECG waveform from PPG alone. However, we argue that this direct mapping is inherently constrained because a single-site peripheral pulse cannot independently capture lead-specific morphological profiles.

Instead, our formulation focuses on reconstructing missing ECG segments by integrating the concurrent PPG signal with the surrounding bidirectional ECG context. This approach is highly clinically relevant because transient ECG signal loss in the NICU typically occurs due to motion artifacts, sensor detachment, or poor electrode contact, meaning neighboring ECG segments and PPG signals often remain available. In this inpainting paradigm, PPG provides complementary hemodynamic timing information, whereas the surrounding ECG context preserves patient-specific and lead-specific morphological information. Therefore, combining these two sources is fundamentally more physiologically plausible and robust than direct full-signal synthesis for reconstructing missing neonatal ECG segments.

### 4.3. Comparison with Previous PPG-to-ECG Reconstruction Studies

Recent studies have demonstrated the feasibility of reconstructing ECG waveforms from PPG signals using diverse deep learning architectures, including U-Net- or convolution-based models [2, 4, 6], Transformer-based models [5], and generative models [21, 22]. Among these studies, PPG2ECGps is particularly relevant to the present work. Tang et al. [20] proposed an end-to-end subject-specific model for reconstructing ECG from PPG without pulse arrival time adjustment. This study provided important evidence that rigid ECG–PPG alignment is not always necessary for PPG-to-ECG reconstruction. Similarly, Transformer-based approaches have recently demonstrated the powerful potential of attention mechanisms for modeling long-range temporal dependencies between PPG and ECG. For example, Lan [5] proposed Performer, which demonstrated the feasibility of reconstructing ECG from adult PPG signals.

However, a critical limitation of these closely related state-of-the-art frameworks is that they were primarily designed for adult data and often evaluated as subject-specific reconstruction frameworks. They did not address neonatal ECG reconstruction under severe random patch loss or continuous block missing conditions. To fully appreciate the structural shift of our model, it is also worth noting broader PPG-to-ECG synthesis attempts. Early approaches attempted to infer ECG morphology using signal-processing or dictionary-learning strategies, while later methods adopted adversarial learning and recurrent architectures. For instance, CardioGAN introduced an attentive generative adversarial framework [21], and subsequent models such as P2E-LGAN explored recurrent temporal modeling [22]. In addition, various U-Net-based and convolutional encoder-decoder architectures have been proposed to capture local temporal patterns [2, 6].

In contrast to these traditional synthesis benchmarks that rely solely on direct PPG-to-ECG mapping, the present study formulates the task as an ECG segment-level inpainting problem for a patient-independent NICU cohort. This distinction is important because PPG alone does not contain lead-specific ECG morphology. By integrating concurrent PPG information with bidirectional ECG context, the proposed architecture achieved stronger morphological preservation than baseline models such as 1D U-Net, TCN, WNet-BiLSTM, and Performer under neonatal missing-signal conditions.

Another important issue in PPG-to-ECG reconstruction is the risk of subject dependency and data leakage. Abdelgaber et al. [3] emphasized that subject-dependent splitting can lead to overly optimistic performance when beats or segments from the same subject appear in both training and testing sets. To address this concern, the present study used a strict patient-independent split, ensuring that all windows from a given patient were assigned exclusively to the training, validation, or test set. This evaluation strategy provides a more realistic estimate of generalization performance in unseen NICU patients.

### 4.4. Physiological Interpretation of Alignment-Free PAT Learning

The temporal relationship between ECG and PPG has traditionally been interpreted using pulse arrival time (PAT) or pulse transit time (PTT), which is commonly measured as the delay between the ECG R-peak and a fiducial point on the PPG waveform [19]. Although PAT and PTT have been widely used for cardiovascular monitoring and cuffless blood pressure estimation, this relationship cannot be fully represented by a single fixed peak-to-peak delay. The ECG–PPG temporal relationship may vary depending on vascular compliance, measurement site, cardiac output, autonomic regulation, and hemodynamic status.

This issue is particularly important in neonates. Premature infants have immature autonomic regulation, high heart rates, narrow QRS complexes, and rapidly changing hemodynamic conditions. Therefore, the temporal delay between ECG and PPG may fluctuate substantially across heartbeats. Under such conditions, artificial alignment based on cross-correlation or fixed fiducial-point matching may impose an oversimplified temporal relationship between the two signals.

Recent pulse-transition studies further support this interpretation. Mohammed et al. [23] showed that pulse-transition features derived not only from ECG R-peaks and PPG rising points but also from ECG T-wave and PPG falling-curve regions can improve non-invasive blood pressure estimation. This finding suggests that physiologically meaningful ECG–PPG coupling may be distributed across broader waveform regions, rather than being confined to a single R-peak-to-PPG fiducial-point pair. Similarly, Abdollahzadeh et al. [24] demonstrated that peak-independent temporal lag modeling can capture hemodynamic dynamics from foot-based PPG without explicit peak detection. Although these studies focused on blood pressure estimation rather than ECG reconstruction, they support the broader concept that temporal delay information between cardiovascular signals can be learned without rigid fiducial-point matching.

The proposed alignment-free architecture was motivated by this physiological consideration. Instead of enforcing artificial synchronization or cross-correlation-based phase correction, the model was allowed to learn temporal correspondences between PPG and masked ECG directly from data. In addition, RoPE was intentionally excluded from the cross-attention module to avoid imposing excessive positional constraints between heterogeneous signals. The attention map analysis provided exploratory evidence that the model utilized physiologically plausible PAT-like temporal information without explicit temporal supervision. This supports the possibility that flexible attention-based coupling may capture neonatal ECG–PPG temporal relationships more appropriately than rigid alignment.

### 4.5. Role of RoPE and Multi-Scale Tokenization

Another important contribution of this study is the integration of relative positional modeling and multi-scale temporal tokenization for neonatal ECG reconstruction. In time-series Transformer models, positional encoding is essential because the temporal order of waveform components directly determines physiological meaning. The original Transformer used absolute positional encoding to inject sequential order information into token representations [12]. However, absolute positional encoding may be suboptimal for physiological signals because the absolute positions of waveform components can shift because of heart rate variability and beat-to-beat temporal variation.

Rotary position embedding (RoPE) encodes positional information through rotations of query and key vectors, allowing attention scores to reflect relative temporal relationships between tokens [13]. This property is suitable for ECG reconstruction because P-QRS-T components preserve relative morphological relationships even when their absolute positions shift. In the proposed model, RoPE was applied to self-attention layers to strengthen internal temporal context modeling within each signal stream. However, RoPE was excluded from cross-attention to avoid forcing a fixed positional correspondence between PPG and ECG. This design was supported by the ablation result showing that applying RoPE to cross-attention reduced reconstruction performance.

The use of multi-scale temporal windows was also motivated by the multi-resolution nature of physiological waveforms. ECG and PPG contain both short-duration morphological features, such as QRS complexes and pulse upstrokes, and longer-range rhythm-level patterns, such as RR intervals and respiratory modulation. Prior time-series Transformer studies have shown that hierarchical multi-scale representations can improve multivariate time-series modeling by capturing both local and global dependencies [25]. This supports the use of multiple temporal window sizes to jointly model rapid waveform transitions and broader rhythm-level context in physiological signal reconstruction.

In the proposed model, short windows were designed to capture rapid waveform transitions related to QRS timing, whereas longer windows were intended to preserve broader cardiac-cycle context and smoother T-wave morphology. We hypothesize that this multi-scale design might have contributed to the model maintaining high component-wise PCC values for QRS, P-wave, and T-wave reconstruction, even under severe random patch and continuous block missing conditions.

### 4.6. Clinical Implications for NICU Monitoring

The clinical motivation of this study is closely related to the limitations of conventional adhesive ECG monitoring in premature infants. Extremely premature infants have immature skin barriers and are vulnerable to epidermal stripping, infection, and MARSI caused by repeated application and removal of adhesive electrodes [1]. Wireless epidermal electronic systems and non-contact physiological monitoring technologies have been proposed to reduce the burden of conventional monitoring in the NICU [15]. However, continuous acquisition of diagnostically useful ECG morphology remains challenging in routine clinical environments.

PPG-based monitoring has gained increasing attention as a less invasive approach for neonatal vital sign monitoring. However, PPG alone cannot provide lead-specific ECG morphology, which is important for rhythm assessment, QRS detection, and waveform interpretation. Therefore, reconstructing missing ECG segments from PPG and surrounding ECG context may provide a practical intermediate solution. The proposed approach does not replace diagnostic ECG monitoring, but it may serve as a signal imputation module during transient signal loss.

The robustness results of the present study support this potential clinical role. The proposed model maintained high morphological similarity under 4.0-second continuous block loss and 60% random patch loss, suggesting that it may help maintain continuous ECG streams during motion artifacts, sensor detachment, or temporary electrode failure. Such reconstructed signals could potentially serve as a basis for exploring downstream clinical applications, such as arrhythmia detection, hypoxemia prediction, hypotension prediction, and early warning models. However, prospective clinical validation is required before the reconstructed ECG can be used for diagnostic decision-making.

### 4.7. Strengths of the Study

This study has several strengths. First, the model was developed using a relatively large NICU cohort and evaluated using a strict patient-independent split, reducing the risk of data leakage and subject-dependent performance inflation. Second, we introduced a premature-infant-tailored adaptive hybrid preprocessing strategy, which improved signal quality while minimizing distortion of sharp QRS complexes that are critical for clinical interpretation. Third, the proposed model used a dual-stream architecture to integrate complementary information from PPG and masked ECG, allowing the model to use both hemodynamic timing and lead-specific ECG context. Fourth, the multi-scale tokenization strategy enabled the model to capture both local waveform morphology and global rhythm patterns. Finally, the intentional exclusion of RoPE from cross-attention provides a useful methodological insight for future medical AI systems that must learn temporal relationships between heterogeneous physiological signals without imposing rigid alignment constraints.

### 4.8. Limitations

Despite its robust performance, this study has several limitations. First, although the proposed model was trained and evaluated using a relatively large NICU dataset, the data were obtained from a single-center cohort. Therefore, multi-center validation is required to assess its generalizability across different monitoring systems, patient populations, and clinical environments. Second, the current model primarily focused on single-lead ECG reconstruction. Further validation is needed to determine whether the proposed framework can be extended to multi-lead ECG reconstruction or spatial vector-related abnormalities. Third, although the model showed robust performance under artificial masking conditions, prospective validation using real-world ECG dropout events is necessary. Artificial masking can simulate clinically relevant signal loss patterns, but it may not fully capture the complexity of actual motion artifacts, electrode detachment, and sensor-related noise in NICU environments. Fourth, the present analysis mainly evaluated reconstruction quality using waveform similarity metrics. Future studies should assess whether reconstructed ECG signals preserve clinically actionable features in arrhythmias, conduction abnormalities, and congenital heart disease-related waveform changes. Fifth, the adaptive preprocessing pipeline used ECG component information during offline signal preparation. Although the target missing segments were not provided to the model after artificial masking, future real-time deployment will require online preprocessing strategies that do not depend on unavailable ECG components within dropout regions. Finally, real-time deployment requires additional engineering validation, including model lightweighting, inference latency measurement, and integration with wireless wearable devices.

### 4.9. Future Directions

Building on the results and limitations of this study, several future directions should be considered. First, multi-center datasets should be established to validate external generalizability and improve robustness across diverse NICU environments. Second, future studies should evaluate the proposed model in pathological ECG patterns, including arrhythmias, conduction abnormalities, and congenital heart disease-related waveform changes. Third, the framework should be extended to multi-lead or spatially informed ECG reconstruction to assess whether bidirectional context referencing can preserve lead-specific morphological information.

Fourth, the reconstructed ECG and synchronized PPG signals may be used to derive hemodynamic features such as PAT, PTT, or pulse wave transit time (PWTT), potentially supporting future non-invasive blood pressure estimation models [19, 24]. By integrating reconstructed high-quality ECG streams with high-resolution vital sign data, this framework could evolve beyond waveform reconstruction into a broader clinical decision support platform for predicting adverse events such as hypoxemia, hypotension, sepsis, or cardiovascular instability.

Finally, prospective clinical trials are needed to evaluate whether a lightweight version of the model can be integrated into wireless wearable monitoring systems and whether such integration can reduce ECG signal loss, decrease electrode replacement frequency, and ultimately reduce MARSI risk in premature infants. Until such validation is completed, the proposed method should be interpreted as a promising signal imputation framework rather than a replacement for standard diagnostic ECG monitoring.

## 5. Conclusion

This study proposed an alignment-free RoPE-based dual-stream Transformer for neonatal ECG segment reconstruction under missing-signal conditions in the NICU. The proposed framework was based on two main concepts: alignment-free PAT-related temporal learning and bidirectional ECG context referencing. By integrating concurrent PPG information with the surrounding ECG context, the model was designed to reconstruct missing

ECG segments without artificial ECG–PPG synchronization.

Using a patient-independent NICU dataset, the proposed model demonstrated high morphological reconstruction performance under a 40% random missing condition (PCC 0.96). In stress-test analyses, it also maintained robust reconstruction performance under a 4.0-second continuous block loss condition (PCC 0.90) and a 60% random patch loss condition (PCC 0.93). These findings suggest that the model can preserve rhythm-level timing and local ECG morphology during transient signal loss.

Nevertheless, this study has several limitations. The dataset was obtained from a single center, and external validation across multiple NICU environments is required. The current framework focused on single-lead ECG reconstruction, and further studies are needed to evaluate its applicability to multi-lead ECG signals and pathological ECG patterns, including arrhythmias and congenital heart disease-related waveform changes. In addition, prospective validation using real-world ECG dropout events and real-time device-level implementation studies are necessary before clinical deployment. Despite these limitations, the proposed framework may serve as a signal imputation module for future non-invasive neonatal monitoring systems. By reducing the impact of transient ECG signal loss, this approach may help maintain ECG monitoring continuity and support downstream clinical decision support systems. However, reconstructed ECG signals should be interpreted as auxiliary signal-recovery outputs rather than replacements for standard diagnostic ECG monitoring until prospective clinical validation is completed.

## Ethics approval

This study was conducted with the approval of the Institutional Review Board of Jeonbuk National University Hospital (IRB No. 2025-12-052-001).

## Declaration of generative AI and AI-assisted technologies in the manuscript preparation process

During the preparation of this work, the authors used ChatGPT to support language editing and improve the clarity of the manuscript. After using this tool, the authors reviewed and edited the content as needed and take full responsibility for the content of the manuscript.

## CRediT authorship contribution statement

SungLim Choi: Conceptualization, Methodology, Software, Formal analysis, Visualization, Writing – original draft. Gyeonmin Gu: Methodology, Validation, Writing – review and editing. YunA Kim: Data curation, Validation, Writing – review and editing. SEUNG HEON Lee: Software, Formal analysis. Sang-Il Sim: Data curation, Resources. Yeong Min Jang: Investigation, Resources. Hyunho Kim: Supervision, Project administration, Clinical interpretation, Writing – review and editing.

## Declaration of competing interest

The authors declare that they have no known competing financial interests or personal relationships that could have appeared to influence the work reported in this paper.

## Data availability

The data used in this study contain sensitive clinical information from NICU patients and are not publicly available due to privacy, ethical, and institutional restrictions. De-identified data may be made available from the corresponding author upon reasonable request and with appropriate institutional approval.

## Notes

### Competing Interest Statement

The authors have declared no competing interest.

### Author Declarations

The Institutional Review Board of Jeonbuk National University Hospital gave ethical approval for this work and waived the requirement for informed consent (IRB No. 2025-12-052-001).

